# Decreased Viral Load, Symptom Reduction, and Prevention of Respiratory Syncytial Virus Infection with MVA-BN-RSV Vaccine

**DOI:** 10.1101/2022.12.02.22283030

**Authors:** Elke Jordan, Golam Kabir, Stephanie Schultz, Günter Silbernagl, Darja Schmidt, Victoria A. Jenkins, Heinz Weidenthaler, Daria Stroukova, Barbara K. Martin, Laurence De Moerlooze

## Abstract

**Background:** Respiratory syncytial virus (RSV) causes significant disease burden in infants and older adults. Most vaccines in development focus on the virus’s F protein. MVA-BN-RSV is a novel vectored vaccine encoding internal and external proteins from both RSV subtypes.

**Methods:** In a phase 2a trial, participants aged 18 to 50 years selected for low RSV titers were randomized to receive MVA-BN-RSV or placebo, then challenged 4 weeks later with RSV-A Memphis 37b. Viral load was assessed from nasal washes and virus cultivation, and RSV symptoms were collected throughout quarantine. Antibody titers and cellular markers were assessed before and after vaccination and challenge.

**Results:** Of 74 participants randomized, 36 received MVA-BN-RSV and 37 received placebo; 31 and 32, respectively, were challenged. Viral load areas under the curve from nasal washes were lower (p=0.017) for MVA-BN-RSV (median=0.00) compared to placebo (median=49.05). Total symptom scores also were lower with MVA-BN-RSV. Vaccine efficacy in preventing infection confirmed by viral culture was 88.5% (CI: 14.8%; 98.5%). Immunoglobulin A and G in serum increased about 4-fold after MVA-BN-RSV vaccination, which was greater than the placebo response to challenge, and neutralizing antibody titer increased about 2-fold. Cellular responses were robust, particularly to the internal RSV proteins. Injection site pain occurred more frequently with MVA-BN-RSV. No serious adverse events were attributed to vaccination.

**Conclusion:** MVA-BN-RSV vaccination resulted in lower viral load and was effective against laboratory-confirmed symptomatic infection. Humoral and cellular responses support broad immunogenicity of the vaccine. No safety issues were identified with vaccination.

**Clinical Trial Registry Number:** NCT04752644

## INTRODUCTION

Respiratory syncytial virus (RSV) remains a common infectious disease without an approved vaccine. It is thought that most children are exposed to RSV by 2 years of age[1,2], and infections can recur throughout life, as protective immunity appears to be incomplete[3]. Infants are understood to be at high risk from RSV; it is estimated that nearly 120,000 children under 5 years of age die from RSV each year globally[4], mostly in the developing world. Older adults are the second highest risk group for severe RSV infection. In high-income countries, an estimated 1.62% of adults ≥60 years of age develop acute RSV infections each year, 0.15% are hospitalized, and approximately 33,000 die of RSV-related causes[5]. Underlying co-morbidities likely drive the risk of severe outcomes in older adults[6,7,8]. Therefore, this population is an important target for preventive measures, as older adults in the developed world may spend decades at risk of both hospitalization and death from RSV.

In the 1960s, research in children with a formalin-inactivated RSV vaccine was halted when it was recognized that those immunized frequently experienced enhanced disease upon later natural infection[9,10,11]. This slowed the pace of RSV vaccine research for decades, as scientists investigated potential mechanisms for this phenomenon[12]. Modern RSV vaccine research has mostly focused on delivery of RSV proteins, rather than whole virus, either as a subunit vaccine or using a viral vector. In particular, vaccine development efforts have aimed to elicit neutralizing antibodies (nAbs) to the surface fusion (F) protein that promotes syncytium formation in respiratory epithelium. Two candidate vaccines for older adults that contained the F protein in its post-fusion conformation were unsuccessful in phase 2 and 3 trials[13,14]. Three other vaccines delivering the F protein in its pre-fusion conformation have reported positive results from phase 2 challenge trials[15,16] and/or from pivotal phase 3 trials[17,18]. None of these vaccines in older adults appears to elicit the imbalanced Th2-mediated immune response thought to be involved in RSV vaccine-enhanced disease[1,2].

MVA-BN-RSV is a novel vaccine aimed at broad immunogenicity, inducing both humoral and cellular responses to multiple RSV proteins. It is a recombinant modified vaccinia Ankara vector that encodes not only the F protein (expressed as both pre- and post-F) but also the surface glycoproteins from the 2 RSV subtypes (G(A) and G(B)) that facilitate viral attachment to airway ciliated epithelial cells and 2 internal proteins, the nucleoprotein (N) and transcription elongation factor (M2-1). The F and G proteins are the main targets of RSV nAbs, but this immune response to natural infection is not durable[3,19]. The N and M2-1 proteins, which are highly conserved among different RSV subtypes, were included in the recombinant vaccine to promote cytotoxic T cell responses.

MVA-BN-RSV may have an advantage over other candidate vaccines that rely on nAbs to a single protein. The vaccine has induced humoral and cellular immune responses in animal models[20] as well as in early clinical trials[21,22] without safety concerns. The safety, efficacy, and immunogenicity of MVA-BN-RSV are examined in this report of a human challenge trial.

## METHODS

### Trial Design, Conduct, and Procedures

#### Design

This was a phase 2a, randomized, double-blind, placebo-controlled trial to assess safety, immunogenicity, and efficacy of MVA-BN-RSV vaccine against infection with the Memphis 37b strain of respiratory syncytial virus subtype A (RSV-A). This virus challenge trial was conducted by hVIVO Services Limited at the Queen Mary BioEnterprises Innovation Centre in London, UK, from January to November 2021.

All trial-related procedures were conducted in accordance with the provisions of the Declaration of Helsinki and approved by the Ethics Committee before trial initiation.

#### Subjects

Subjects provided written informed consent before participating in the trial. They were healthy adults between 18 and 50 years of age expected to be susceptible to RSV based on their nAb titers at screening. Full eligibility criteria are provided in the Supplemental Materials.

#### Vaccination Phase

The MVA-BN-RSV vaccine is based on the modified vaccinia Ankara vector and genetically engineered to encode the RSV F, G(A), G(B), N, and M2-1 proteins. The vaccine was produced at Bavarian Nordic A/S (Kvistgård, Denmark) according to GMP standards. Subjects were randomized 1:1 to be vaccinated intramuscularly with a nominal titer of 5 × 10E8 infectious units per 0.5 mL dose or an equal volume of Tris buffered saline solution. Subjects used a memory aid card to collect information on local (pain, erythema, swelling, induration, and pruritus) and systemic (pyrexia, headache, myalgia, chills, nausea, and fatigue) reactions on the day of and for 7 days after vaccination. Blood was collected for antibody titers and cellular markers before vaccination, 7 days after vaccination for cellular markers, and 14 days after vaccination for antibody titers.

#### Challenge and Quarantine Phase

Use of the human challenge model in RSV research has been well described[23,24]. RSV-A Memphis 37b at a dose of 4.5 log_10_ plaque-forming units was utilized as the challenge virus. Subjects were admitted to a quarantine unit 2 days before being inoculated intranasally with the challenge virus (approximately 4 weeks after vaccination) and were observed for 12 days following challenge. Subjects rated their experience of 13 symptoms on a diary card twice on the first day of quarantine, once on the last day, and 3 times each day in between. Most symptoms were scored in severity from 0 to 3; shortness of breath and wheezing had a fourth severity category for symptoms at rest. Details of symptom scoring is provided in the Supplementary Materials. Nasal washes were obtained twice a day from the second to eleventh day after challenge and once on the final quarantine day. Blood was collected for antibody titers and cellular markers before challenge and 5 and 10 days after challenge.

#### Follow-up Phase

Subjects were followed after discharge from quarantine, and blood was collected for antibody titers and cellular markers at 4 weeks after challenge and approximately 6 months after vaccination.

#### Entire Trial Period

Reports of unsolicited adverse events (AEs) were collected throughout the trial, from the time of informed consent to trial end, and followed until resolution.

### Laboratory Measures

#### Viral Load

A validated reverse transcriptase quantitative polymerase chain reaction (qRT-PCR) was used both to determine whether RSV-A Memphis 37b could be detected and to measure the amount of challenge virus present (lower limit of quantitation [LLOQ] was defined as a cycle threshold [Ct] value of 3.9, which equated to 2.8 log_10_ copies/mL) in nasal washes. Results below the LLOQ were set to 0. Nasal washes also were cultured for replicating virus, and the results were measured by plaque assay; the LLOQ was 2.0 log_10_ plaque-forming units (PFU)/mL, and results below the LLOQ were set to 0. Refer to Supplementary Materials for details.

#### Immunogenicity Measures

Serum samples were analyzed by enzyme-linked immunosorbent assay (ELISA) for titers of RSV-specific immunoglobulins A (IgA) and G (IgG) and by plaque reduction neutralization test (PRNT) for neutralizing titers of RSV Subtype A- and Subtype B-specific antibodies. A double-color enzyme-linked immunospot (ELISpot) was used to enumerate peripheral blood mononuclear cells (PBMCs) producing interferon-γ (IFN-γ) and interleukin-4 (IL-4) in response to stimulating pools representing the surface proteins F, G(A), and G(B); the internal proteins M2-1 and N; and whole RSV. These methods have been previously described[21,22].

### Incidence of Infection and Vaccine Efficacy

Incidence by qRT-PCR in each treatment group was calculated as the proportion of subjects infected from Day 2 to end of quarantine, according to 3 a priori definitions: 1) infection confirmed by qRT-PCR with RSV detectable in samples from at least 2 consecutive days AND symptomatic as evidenced by one or more clinical symptoms of grade ≥2; 2) detectable infection with symptoms as above or with 1 or more symptoms of any grade from 2 different categories in the symptoms scoring system; or 3) detectable infection regardless of symptoms. Vaccine efficacy was defined as (1 – incidence ratio) × 100%.

Post hoc, it was recognized that other recently published human challenge trials[15,16] included definitions of infection based on quantifiable (rather than detectable) qRT-PCR measures, as detectable measures were too sensitive, and respiratory symptoms did not add sufficient specificity to the definitions of infection. Definitions therefore were expanded to include corollary definitions based on quantifiable qRT-PCR measures. Additionally, p-values and estimates of vaccine efficacy were calculated for the exploratory endpoints of infection incidence based on quantifiable virus culture results.

### Statistical Analysis

All statistical analyses were performed using SAS 9.4 (SAS-Institute, Cary, NC, USA). The data were analyzed by Venn Life Sciences.

The primary and secondary efficacy analyses were performed in the per protocol (PP) population, which included all participants who were vaccinated, challenged, and had nasal washes at least until Day 10 of quarantine. The intent-to-treat (challenge) (ITTc) population, which included all vaccinated and challenged participants, was used for supportive analyses on efficacy endpoints. Safety endpoints were analyzed on all vaccinated participants (safety population).

Summary statistics were calculated for demographic and baseline characteristics and efficacy and safety endpoints. For viral load area under the curve (AUC), confirmatory testing of treatment differences was done with a 1-sided Wilcoxon rank sum test. Descriptive treatment comparison of peak viral load, quantitative viral culture, peak viral culture, sum of total symptom scores (TSSs), and peak TSS were performed with 2-sided Wilcoxon rank sum tests. For the incidence of RSV-A Memphis 37b infection detected by qRT-PCR and quantified by qRT-PCR and incidence of infection confirmed by virus culture, descriptive treatment comparisons were performed using Fisher’s exact test. The Wilson score method was used to calculate confidence intervals for proportions. The duration of viral shedding, defined as the time from first detectable viral load until the time after which no further virus was detected, was analyzed by the Kaplan-Meier method. A stepwise logistic regression was performed to explore pre-challenge immune markers prognostic of symptomatic RSV infection.

For safety data, unsolicited adverse events were coded using the Medical Dictionary for Regulatory Activities, version 24.0.

## RESULTS

### Participant Demographics and Characteristics

A total of 74 participants were randomized, 36 to receive MVA-BN-RSV and 38 to receive placebo in a blinded fashion. All but one were vaccinated and are included in the safety analysis set. Ten participants, 5 from each treatment group, did not proceed with RSV-A Memphis 37b inoculation. Of the 63 participants inoculated (ITTc analysis set), 1 participant in each group did not have viral load samples collected through Day 10 (study withdrawals); therefore 61 participants are included in the PP analysis set. Participant disposition is represented in Figure 1.

**Figure 1:**
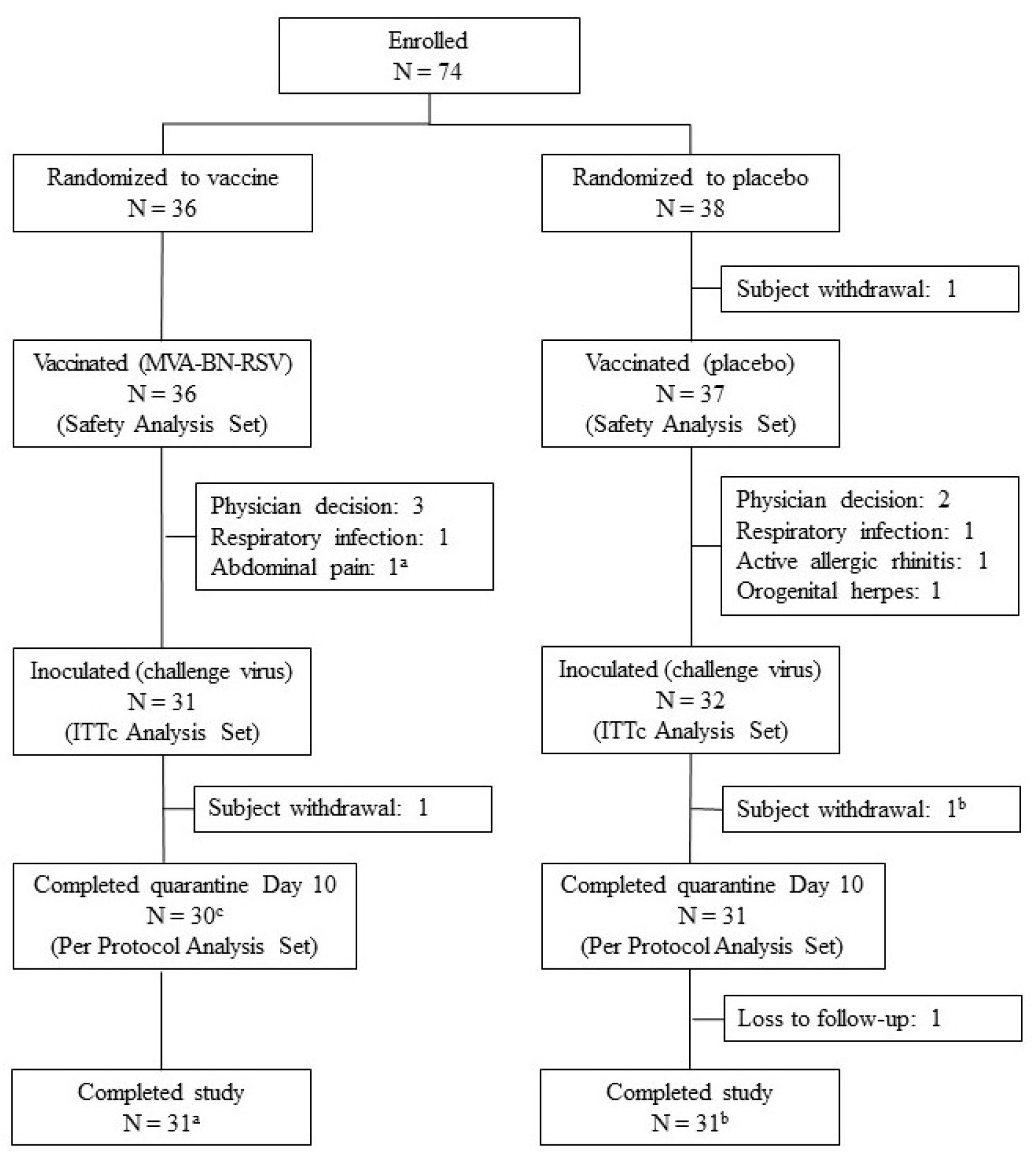
Subject Disposition (All Participants) ITTc = intent-to-treat (challenge); N = number of participants; PP = per protocol. ^a^ Subject with abdominal pain was not inoculated but returned for the final study visit. Subject is not counted in ITTc or PP analysis sets but is recorded as completing the study. ^b^ Subject with myocarditis withdrew before Day 10 of quarantine was completed but returned for the follow-up visits. Subject is not counted in PP analysis set but is recorded as completing the study. ^c^ Subject with myocarditis discontinued after Day 10 of quarantine and returned for the follow-up visits. Subject is counted in PP analysis set and recorded as completing the study.

The baseline characteristics (Table S1) and medical history of the participants were similar between the MVA-BN-RSV and placebo groups.

### Efficacy Results

As shown in Table 1, the primary endpoint of RSV-A Memphis 37b viral load AUC (log_10_ copies×hour/mL) from nasal washes as determined by qRT-PCR was lower in the MVA-BN-RSV group (median=0.00, interquartile range 0.00 to 53.44) than in the placebo group (median=49.05, interquartile range 0.00 to 999.94) (p=0.017). Mean viral load over time is shown in Figure 2A; it diverged 2 days after challenge and remained divergent for all of quarantine. Similarly, viral load AUC by quantitative virus culture was lower in the MVA-BN-RSV group (p<0.001); Figure 2B shows that this difference occurred primarily from Day 3 to Day 7. In addition to viral load AUCs, which were measures of disease summed over time, peak viral load measures by qRT-PCR demonstrated less disease acuity in the MVA-BN-RSV group (median=0.00) than in the placebo group (median=3.45) (p=0.032). The picture was much the same for clinical symptoms; both symptoms summed over time (see Figure 2C) and peak symptom scores were lower in those vaccinated with MVA-BN-RSV, with symptom scores diverging Day 4 through Day 9.

**Table 1:**
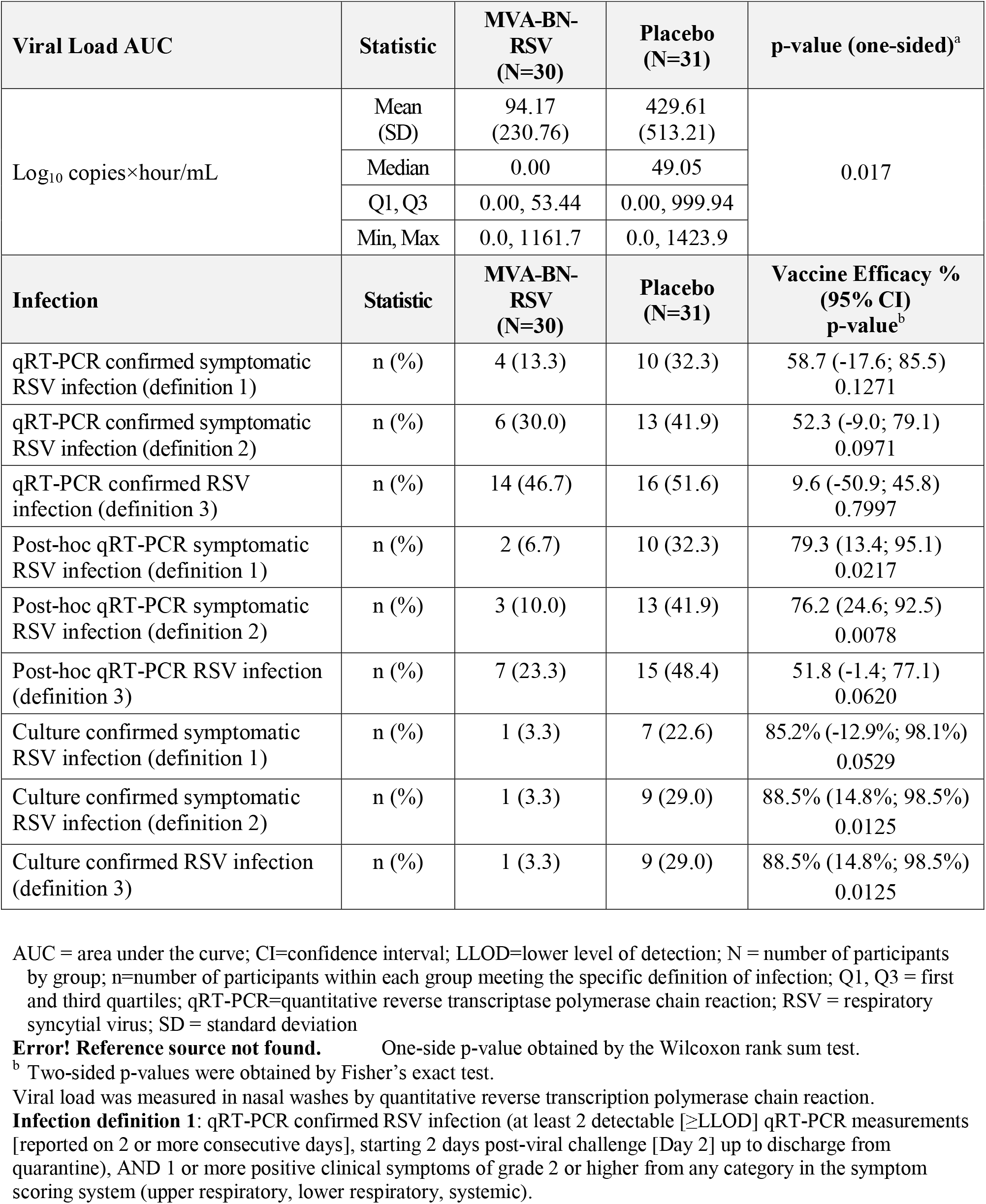
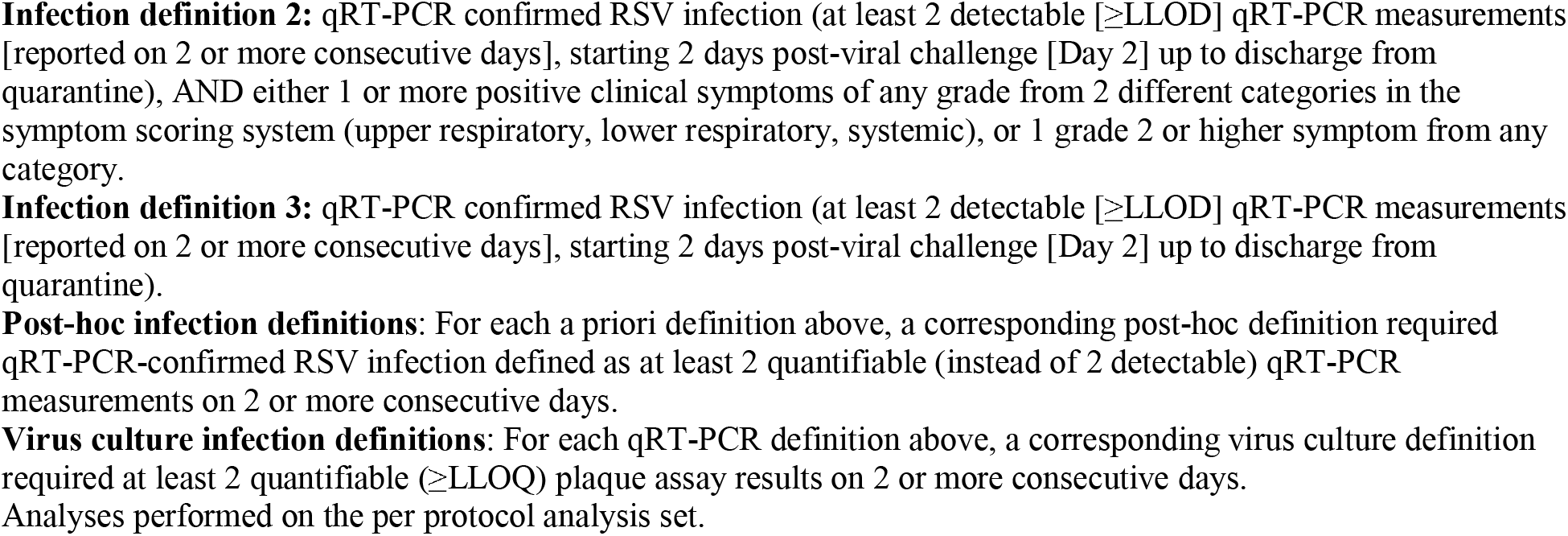
RSV-A Memphis 37b Viral Load Area Under the Curve, Incidence of Infection, and Vaccine Efficacy.

| Viral Load AUC | Statistic | MVA-BN-RSV<br>(N=30) | Placebo<br>(N=31) | p-value (one-sided)<sup>a</sup> |
| --- | --- | --- | --- | --- |
| Log<sub>10</sub> copies×hour/mL | Mean<br>(SD) | 94.17<br>(230.76) | 429.61<br>(513.21) | 0.017 |
|  | Median | 0.00 | 49.05 |  |
|  | Q1, Q3 | 0.00, 53.44 | 0.00, 999.94 |  |
|  | Min, Max | 0.0, 1161.7 | 0.0, 1423.9 |  |
| Infection | Statistic | MVA-BN-RSV<br>(N=30) | Placebo<br>(N=31) | Vaccine Efficacy %<br>(95% CI)<br>p-value<sup>b</sup> |
| qRT-PCR confirmed symptomatic RSV infection (definition 1) | n (%) | 4 (13.3) | 10 (32.3) | 58.7 (-17.6; 85.5)<br>0.1271 |
| qRT-PCR confirmed symptomatic RSV infection (definition 2) | n (%) | 6 (30.0) | 13 (41.9) | 52.3 (-9.0; 79.1)<br>0.0971 |
| qRT-PCR confirmed RSV infection (definition 3) | n (%) | 14 (46.7) | 16 (51.6) | 9.6 (-50.9; 45.8)<br>0.7997 |
| Post-hoc qRT-PCR symptomatic RSV infection (definition 1) | n (%) | 2 (6.7) | 10 (32.3) | 79.3 (13.4; 95.1)<br>0.0217 |
| Post-hoc qRT-PCR symptomatic RSV infection (definition 2) | n (%) | 3 (10.0) | 13 (41.9) | 76.2 (24.6; 92.5)<br>0.0078 |
| Post-hoc qRT-PCR RSV infection (definition 3) | n (%) | 7 (23.3) | 15 (48.4) | 51.8 (-1.4; 77.1)<br>0.0620 |
| Culture confirmed symptomatic RSV infection (definition 1) | n (%) | 1 (3.3) | 7 (22.6) | 85.2% (-12.9%; 98.1%)<br>0.0529 |
| Culture confirmed symptomatic RSV infection (definition 2) | n (%) | 1 (3.3) | 9 (29.0) | 88.5% (14.8%; 98.5%)<br>0.0125 |
| Culture confirmed RSV infection (definition 3) | n (%) | 1 (3.3) | 9 (29.0) | 88.5% (14.8%; 98.5%)<br>0.0125 |

AUC = area under the curve; CI=confidence interval; LLOD=lower level of detection; N = number of participants by group; n=number of participants within each group meeting the specific definition of infection; Q1, Q3 = first and third quartiles; qRT-PCR=quantitative reverse transcriptase polymerase chain reaction; RSV = respiratory syncytial virus; SD = standard deviation
**Error! Reference source not found.** One-side p-value obtained by the Wilcoxon rank sum test.
<sup>b</sup> Two-sided p-values were obtained by Fisher's exact test.
Viral load was measured in nasal washes by quantitative reverse transcription polymerase chain reaction.
**Infection definition 1:** qRT-PCR confirmed RSV infection (at least 2 detectable [ ≥ LLOD] qRT-PCR measurements [reported on 2 or more consecutive days], starting 2 days post-viral challenge [Day 2] up to discharge from quarantine), AND 1 or more positive clinical symptoms of grade 2 or higher from any category in the symptom scoring system (upper respiratory, lower respiratory, systemic).
**Infection definition 2:** qRT-PCR confirmed RSV infection (at least 2 detectable [ ≥ LLOD] qRT-PCR measurements [reported on 2 or more consecutive days], starting 2 days post-viral challenge [Day 2] up to discharge from quarantine), AND either 1 or more positive clinical symptoms of any grade from 2 different categories in the symptom scoring system (upper respiratory, lower respiratory, systemic), or 1 grade 2 or higher symptom from any category.
**Infection definition 3:** qRT-PCR confirmed RSV infection (at least 2 detectable [ ≥ LLOD] qRT-PCR measurements [reported on 2 or more consecutive days], starting 2 days post-viral challenge [Day 2] up to discharge from quarantine).
**Post-hoc infection definitions:** For each a priori definition above, a corresponding post-hoc definition required qRT-PCR-confirmed RSV infection defined as at least 2 quantifiable (instead of 2 detectable) qRT-PCR measurements on 2 or more consecutive days.
**Virus culture infection definitions:** For each qRT-PCR definition above, a corresponding virus culture definition required at least 2 quantifiable ( ≥ LLOQ) plaque assay results on 2 or more consecutive days. Analyses performed on the per protocol analysis set.

**Figure 2:**
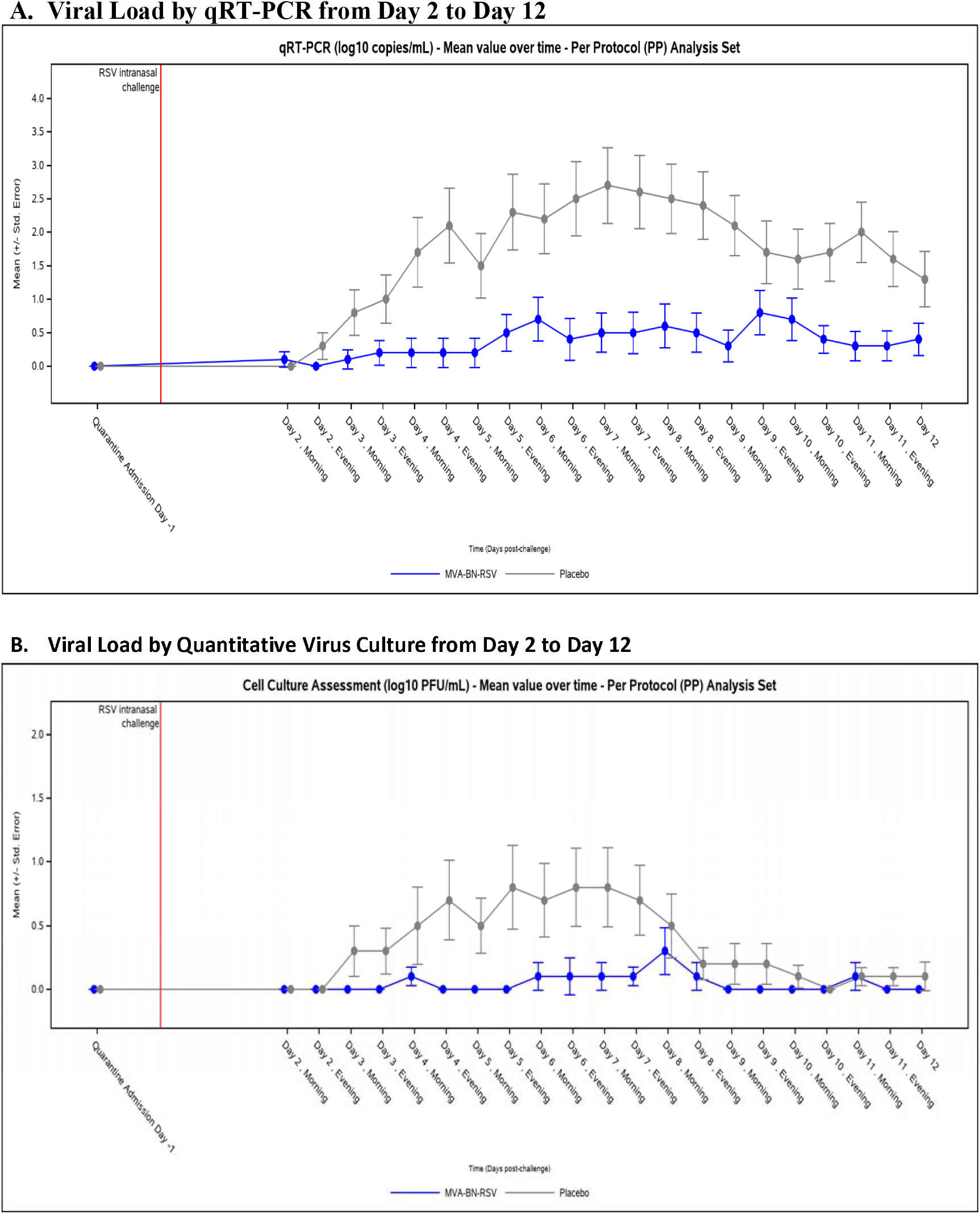
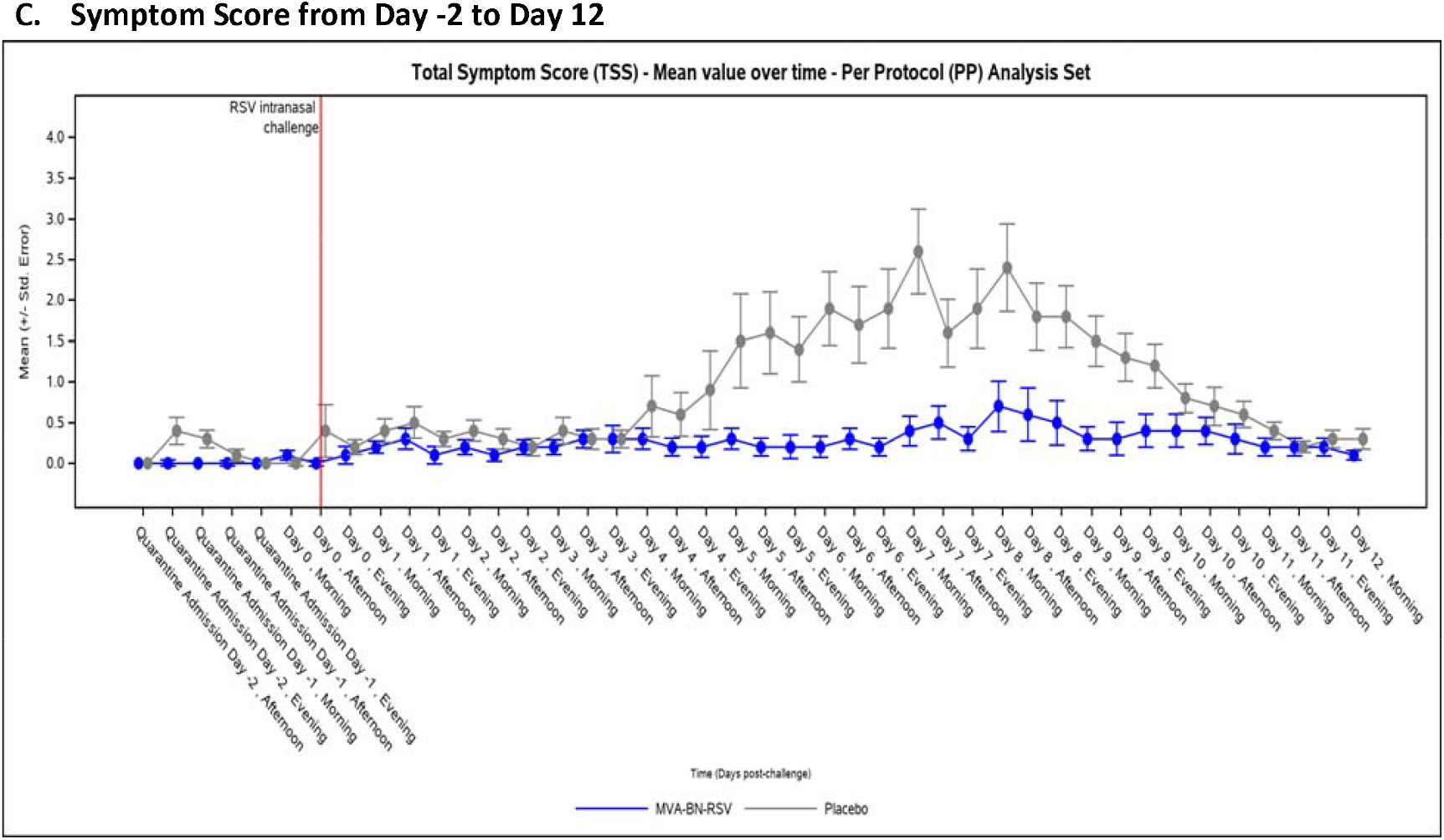
Mean Viral Load and Symptoms Over Time. PFU = plaque-forming units; PP = per protocol; qRT-PCR = quantitative reverse transcription polymerase chain reaction; RSV = respiratory syncytial virus; TSS = total symptom score. X axis values are slightly offset for better readability of the comparison between treatment groups. Analyses performed on the per protocol analysis set. Viral load results that were less than the lower limit of quantification were arbitrarily set to 0.

Incidence of RSV-A Memphis 37b infection was investigated using several definitions, as described in the methods, and the results are presented in Table 1. Under the a priori definitions based upon detectable qRT-PCR viral load, vaccine efficacy ranged from a low of 9.6% for infection confirmed by laboratory measure only to 58.7% for infection confirmed both by laboratory measure and by the presence of at least one RSV symptom of grade ≥2, and the differences in infection incidence between the treatment groups were not statistically significant. When infection was defined instead by quantifiable qRT-PCR viral load, efficacy ranged from 51.8% to 79.3%, and differences in symptomatic infection were statistically significant. Finally, when defined by virus culture results alone, vaccine efficacy was 88.5% (p=0.0125); addition of clinical symptoms to the definition did not improve efficacy.

The exploratory outcome of duration of viral shedding is presented in Figure 5A. Viral shedding for the MVA-BN-RSV group, when it occurred, mostly ended by Day 4; one participant was not clear of viral shedding until Day 8. For the placebo group, viral shedding was mostly sustained to Day 6 and longer.

### Immunogenicity Results

RSV-specific humoral responses are shown in Figure 3A to 3D and Table S2. In the MVA-BN-RSV group, the largest increases were seen in IgA and IgG GMTs, which increased about 4-fold at 2 weeks after vaccination; neither immunoglobulin increased in response to challenge. The placebo group experienced over 2-fold increases in IgA and IgG in response to challenge; these responses were slower than the MVA-BN-RSV group’s responses after vaccination, as they peaked at 28 days after challenge. The MVA-BN-RSV group had higher IgG and IgA GMTs than the placebo group at the end of the study, but both groups remained above baseline. For PRNT-A and PRNT-B, the MVA-BN-RSV group experienced about a 2-fold increase at 2 weeks after vaccination; they had no response to challenge. The placebo group’s response to challenge was slower and peaked at higher levels at 28 days after challenge compared to the MVA-BN-RSV group’s peak following vaccination. Both groups’ nAb titers were greater than baseline at the end of the study.

**Figure 3:**
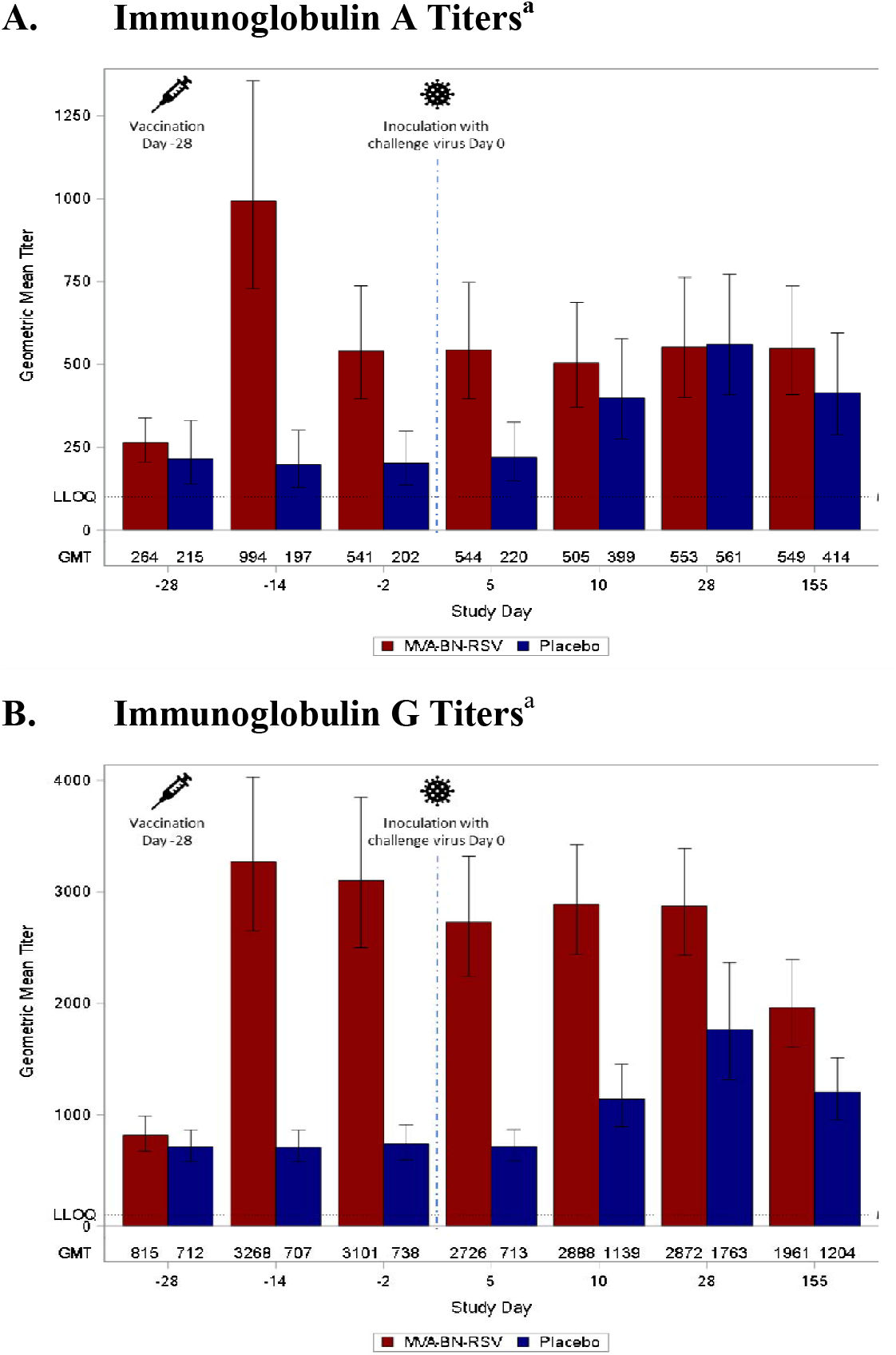
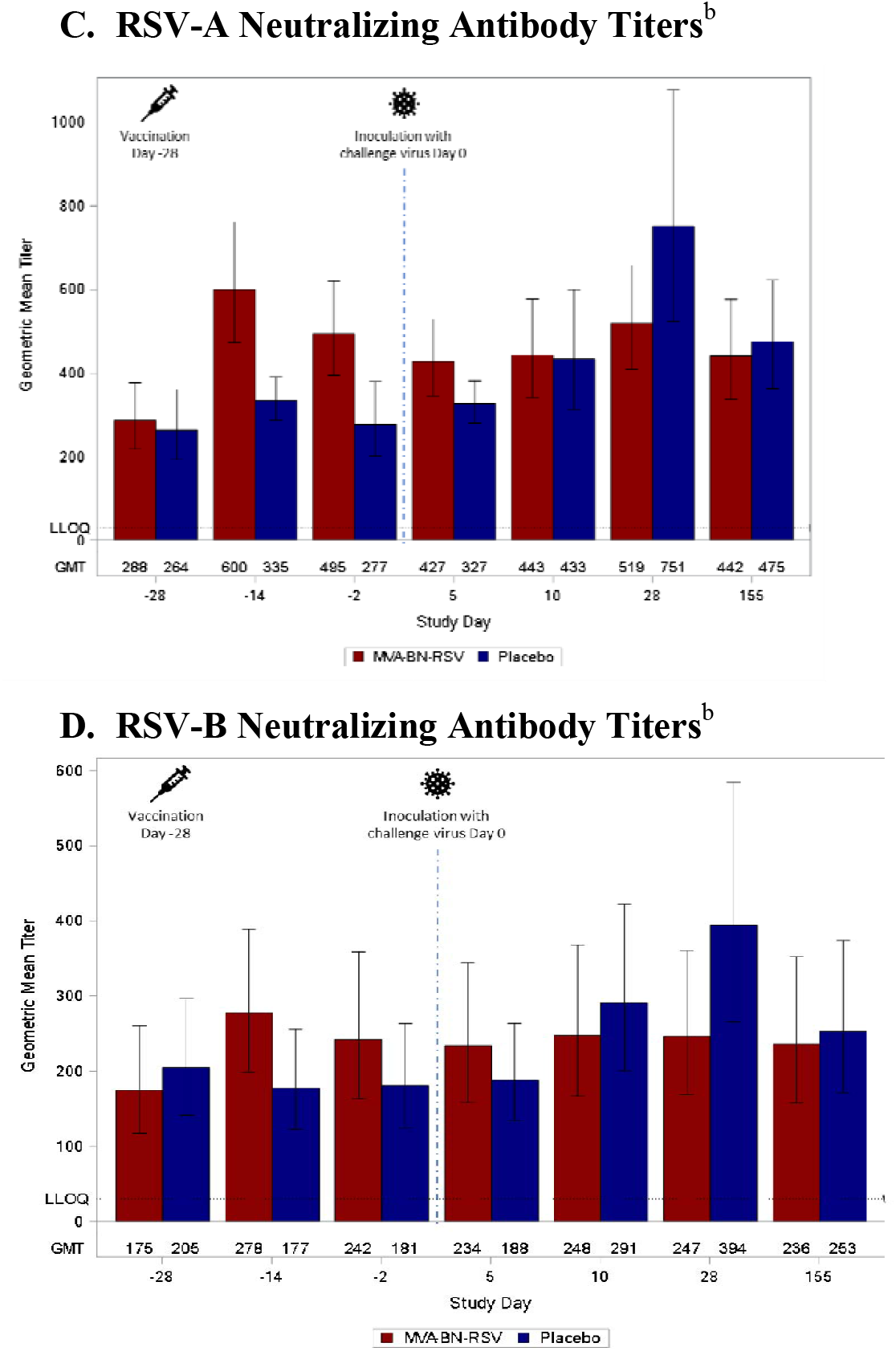
RSV-specific Antibodies. GMT = geometric mean titer; IU = infectious units; LLOQ = lower limit of quantification; RSV = respiratory syncytial virus. ^a^ Immunoglobulin titers were analyzed by enzyme-linked immunosorbent assay. ^b^ Neutralizing antibody titers were analyzed by plaque reduction neutralization test, standardized to World Health Organization standard as IU/mL. Day -28 is baseline (measures obtained on the day of and just prior to vaccination), Day -14 captures vaccine effects 2 weeks after vaccination, and Day -2 is pre-inoculation. Inoculation occurred on Day 0. Days 5, 10, and 28 capture effects after inoculation, and Day 155 represents follow-up approximately 6 months after vaccination. The results <LLOQ are considered by using ½LLOQ for the calculation of statistics for the various antibody types. Analyses performed on the per protocol analysis set.

For IFN-γ-producing PBMCs (Figure 4 and Table S3), geometric mean spot-forming units (GMSFUs) increased 2- to 6-fold in response to the various stimulating pools by 7 days after vaccination with MVA-BN-RSV, and the increase was generally larger than that seen in the placebo group at peak (10 days after inoculation with the challenge virus). Responses to the M2-1 and N pools remained greater than twice the baseline levels for the MVA-BN-RSV group at the end of the study. By contrast, GMSFUs for the placebo group returned to baseline or just slightly above for all pools. GMSFUs for IL4-producing PBMCs were low throughout the study (data not shown), suggesting a cellular response favoring Th1 rather than Th2.

**Figure 4:**
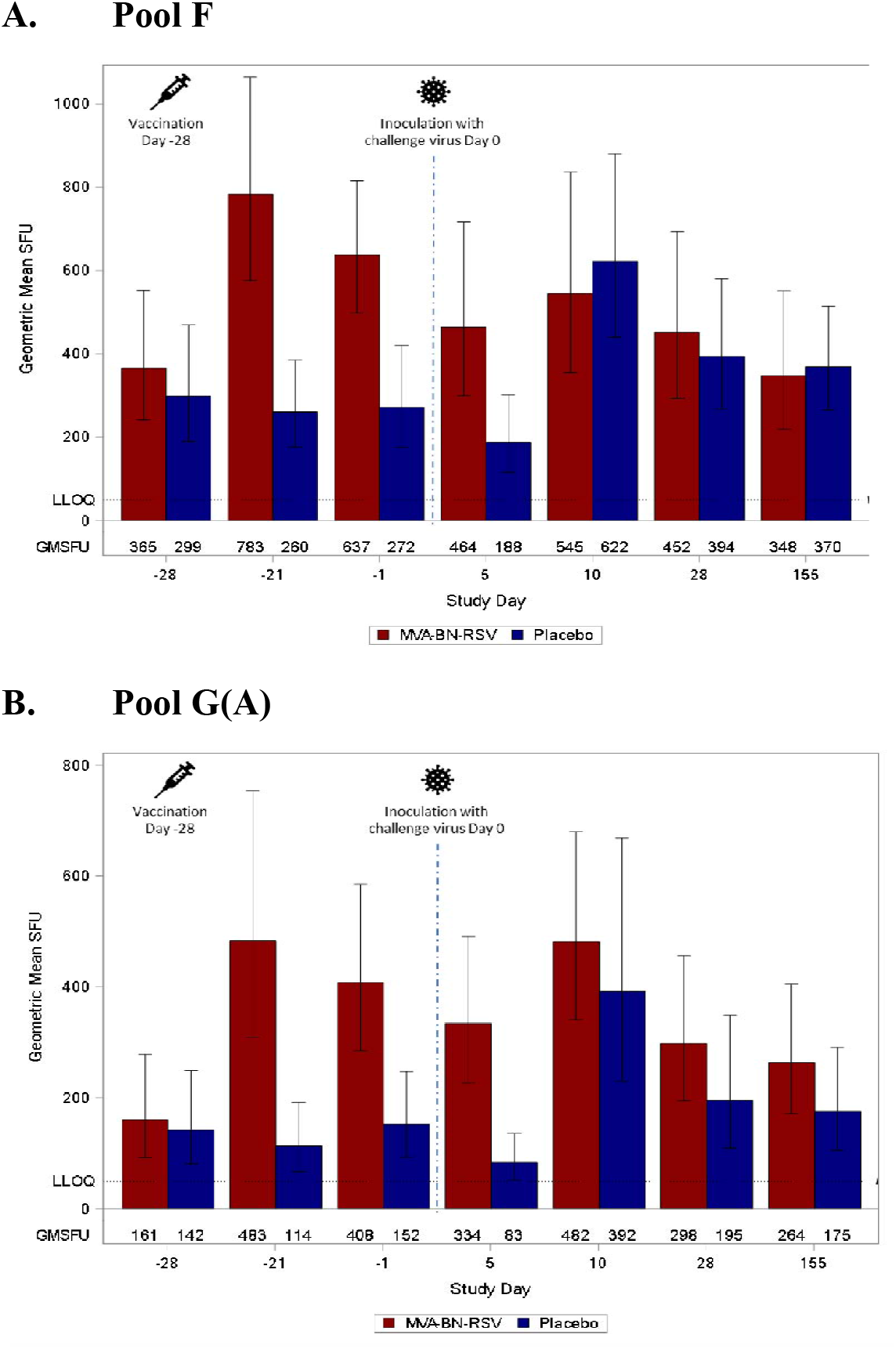
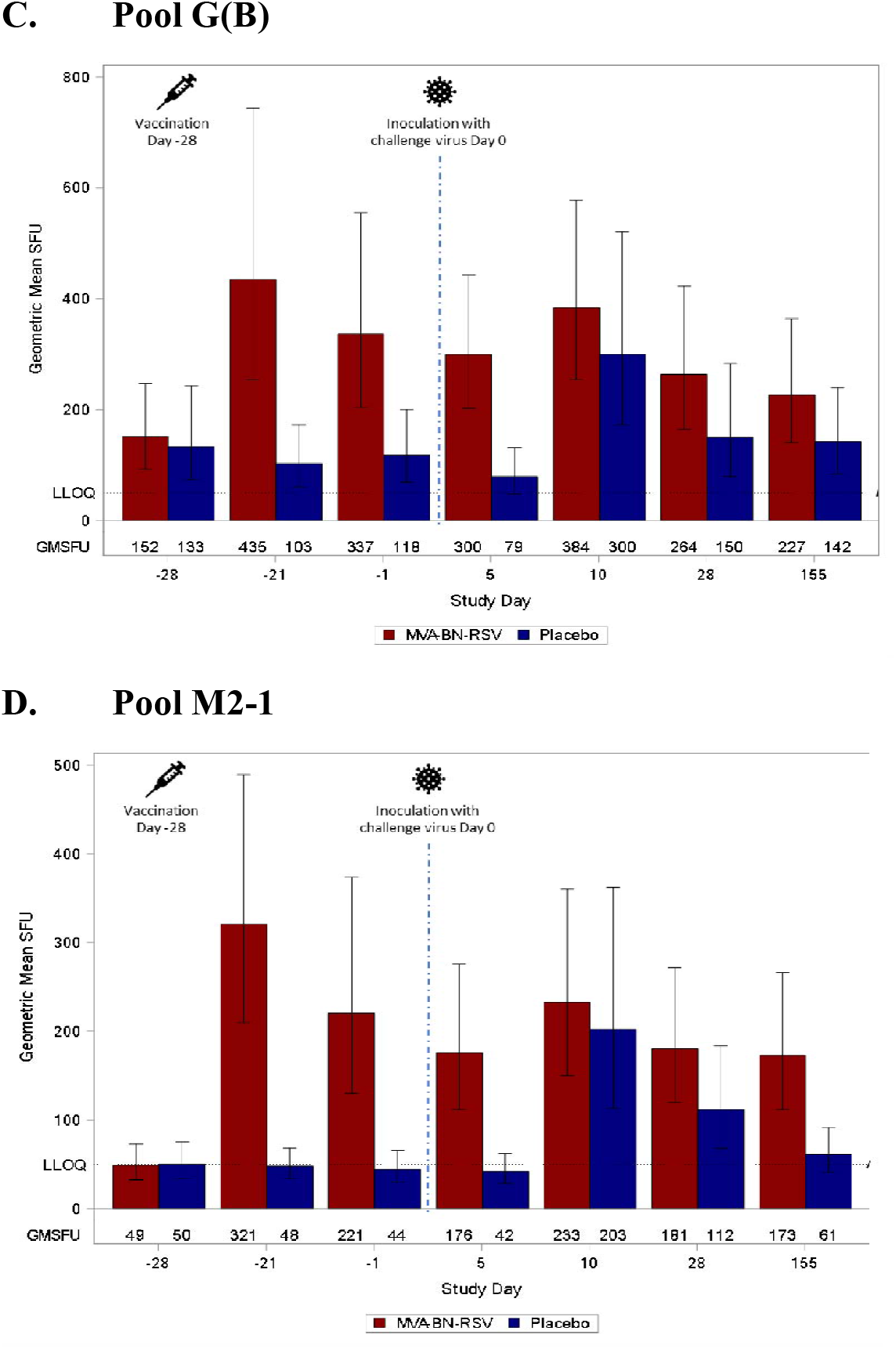
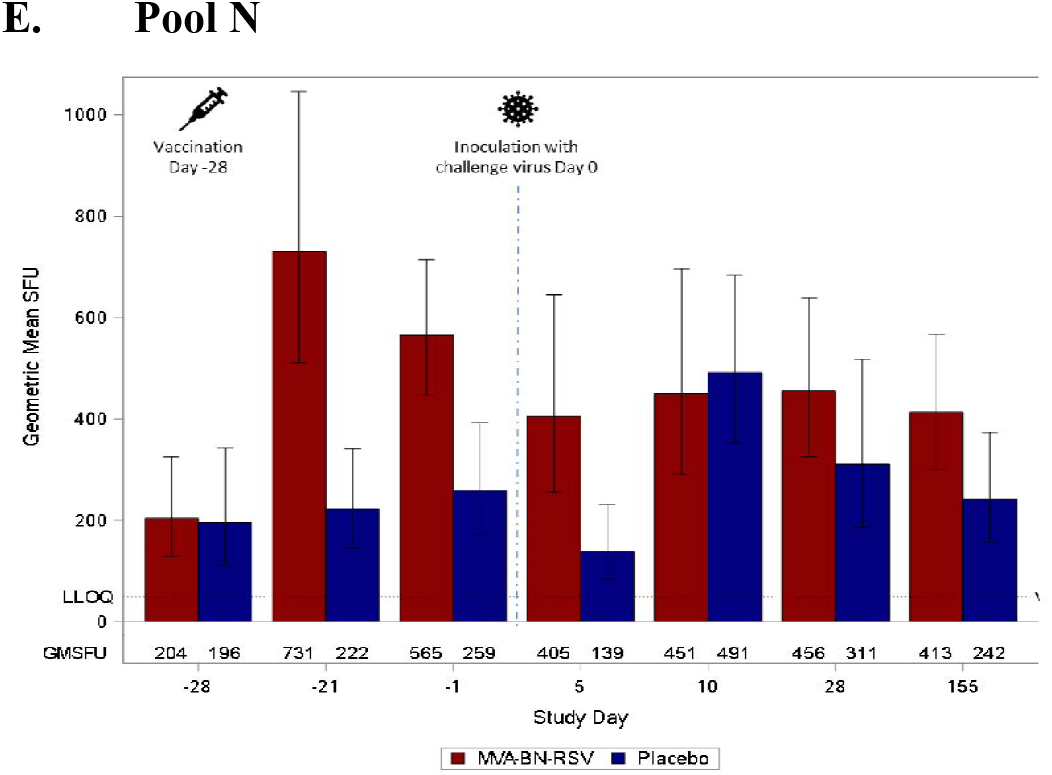
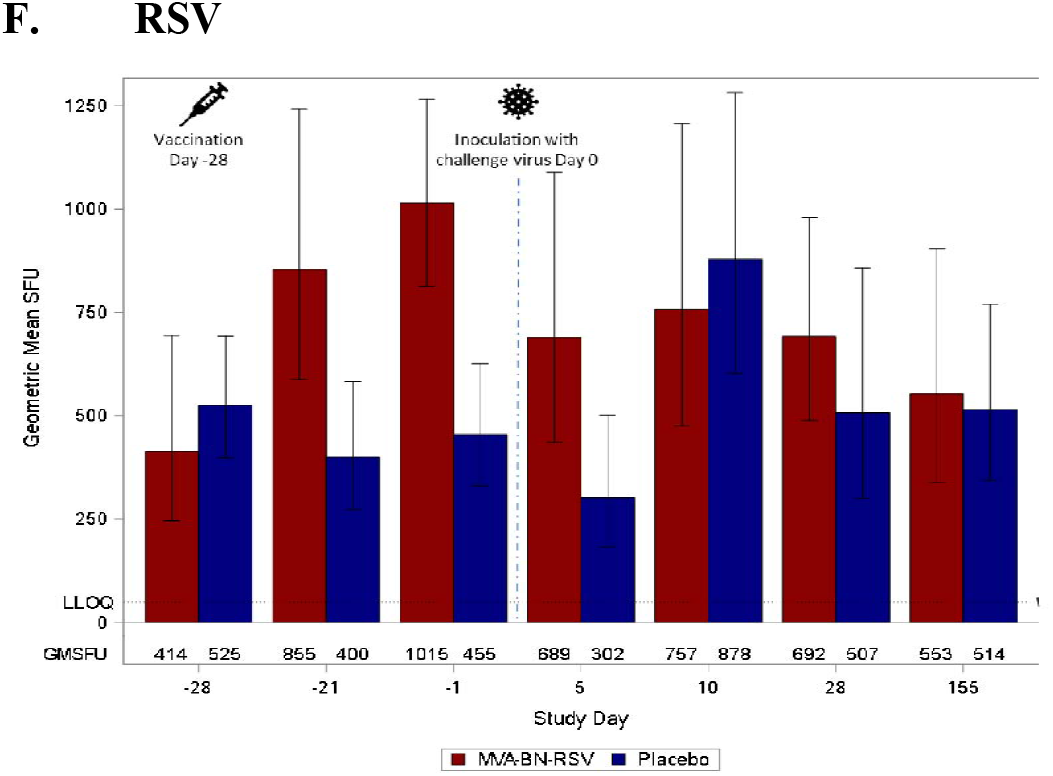
RSV-specific Cellular Responses: Enumeration of Interferon-γ-producing Peripheral Blood Mononuclear Cells. GMSFU = geometric mean spot forming units; LLOQ = lower limit of quantification; RSV = respiratory syncytial virus. Day -28 is baseline (measures obtained on the day of and just prior to vaccination), Day -14 captures vaccine effects 2 weeks after vaccination, and Day -2 is pre-inoculation. Inoculation occurred on Day 0. Days 5, 10, and 28 capture effects after inoculation, and Day 155 represents follow-up approximately 6 months after vaccination. Enumeration of interferon-γ-producing peripheral mononuclear blood cells was analyzed by enzyme-linked immunosorbent spot assay. The results <LLOQ are considered by using ½LLOQ for the calculation of statistics for the various pools. Analyses performed on the per protocol analysis set.

**Figure 5:**
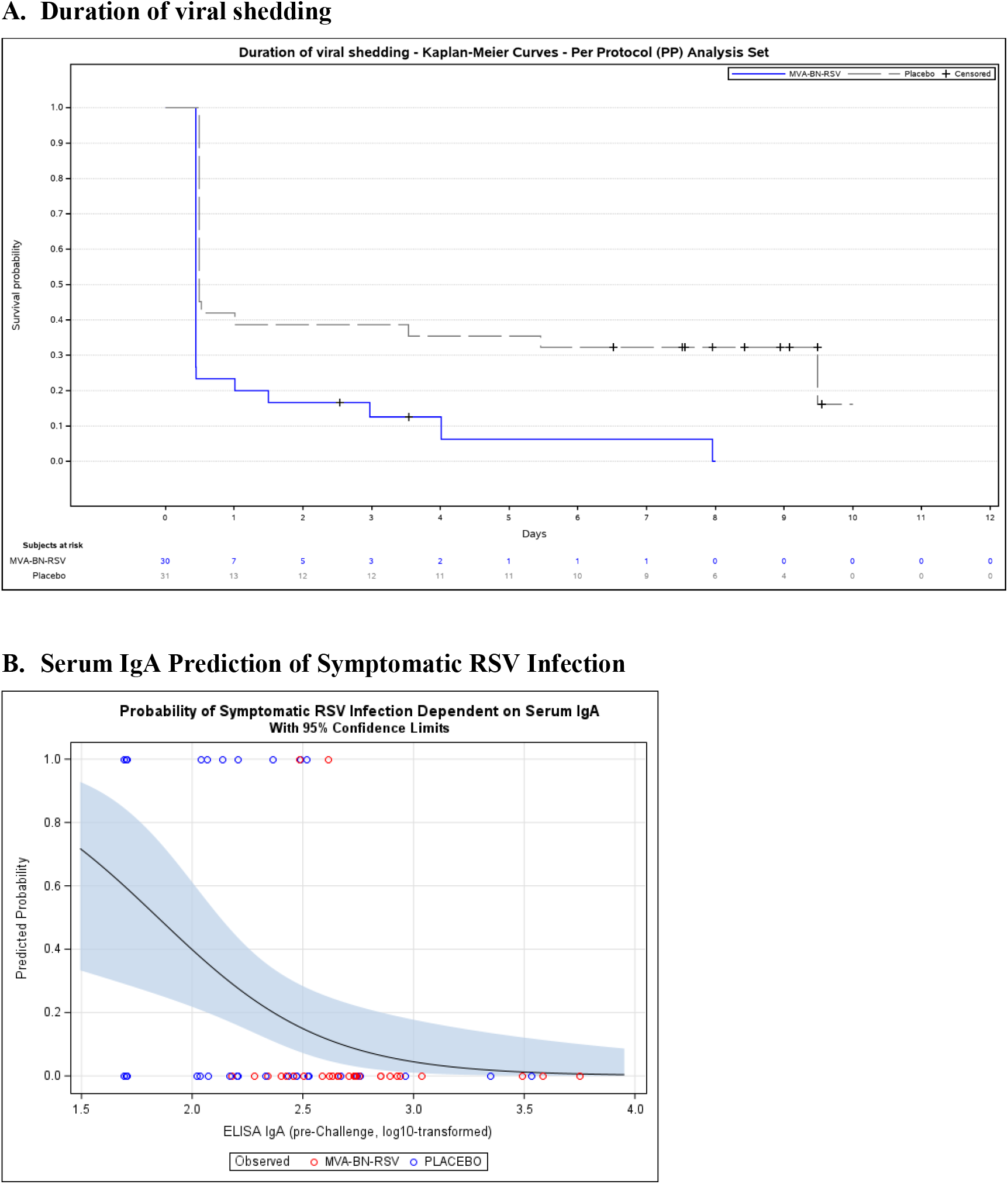
Viral Shedding and Prediction of Infection. ELISA = enzyme-linked immunosorbent assay; IgA = immunoglobulin A; RSV = respiratory syncytial virus. Duration of viral shedding was defined as the time from first detectable viral load until the time after which no further virus was detected. Analyses performed on the per protocol analysis set.

Figure 5B displays the predicted probabilities for symptomatic RSV infection as a function of pre-challenge IgA serum values, the strongest prognostic factor (p=0.020), with higher values being more protective.

### Safety Results

Solicited local AEs (Table 2) were reported quite frequently in the MVA-BN-RSV group; the most common local reaction was injection site pain, and 3 participants reported grade 3 pain. The median duration of pain in the MVA-BN-RSV group was 4.0 days; the maximum was 7 days. Solicited systemic AEs were also common but were reported somewhat more frequently than local reactions by the placebo group as well. Grade 3 fatigue was reported by 4 MVA-BN-RSV recipients, one of whom also reported grade 3 myalgia, and by 1 placebo recipient. The median duration of all systemic AEs was 1.0 days. Unsolicited AEs in the vaccination phase (within 29 days post vaccination) were reported by a comparable proportion of participants in the MVA-BN-RSV and placebo groups. Most were considered unrelated to study vaccination, though 1 case of fatigue and increased body temperature in the MVA-BN-RSV group and 1 case of pruritic rash in the placebo group were attributed to vaccination. Unsolicited AEs in the post-inoculation phase also were reported by similar proportions of the MVA-BN-RSV and placebo groups. No events were considered related to study vaccination. Two SAEs of myocarditis (1 event of moderate severity in the MVA-BN-RSV group that started 10 days after challenge and 1 event of mild severity in the placebo group that started 7 days after challenge) were considered probably related to RSV-A Memphis 37b inoculation. Both were diagnosed by a cardiologist from electrocardiogram and cardiac enzyme findings with no or minimal symptoms and were sent to hospital for observation and follow-up. Tests returned to normal, and the subjects completed follow-up.

**Table 2:** Treatment-emergent Adverse Events.

| **Adverse Events**<br>n (%) [events] | **MVA-BN-RSV** | **Placebo** |
| --- | --- | --- |
| **Vaccinated subjects** | **N=36** | **N=37** |
| **Solicited local AEs** |  |  |
| Any solicited local AE<sup>a</sup> | 32 (88.9) [60] | 14 (37.8) [17] |
| Pain | 30 (83.3) [30] | 4 (10.8) [4] |
| Erythema | 16 (44.4) [16] | 8 (21.6) [8] |
| Induration | 7 (19.4) [7] | 0 |
| Swelling | 5 (13.9) [5] | 3 (8.1) [3] |
| Pruritis | 2 (5.6) [2] | 2 (5.4) [2] |
| Any solicited local AE, grade $\geq 3$ | 3 (8.3) [3] | 0 |
| **Solicited systemic AEs** |  |  |
| Any solicited systemic AE<sup>b</sup> | 29 (80.6) [68] | 19 (51.4) [29] |
| Fatigue | 17 (47.2) [17] | 12 (32.4) [12] |
| Myalgia | 21 (58.3) [21] | 4 (10.8) [4] |
| Headache | 15 (41.7) [15] | 9 (24.3) [9] |
| Chills | 8 (22.2) [8] | 1 (2.7) [1] |
| Pyrexia | 6 (16.7) [6] | 0 |
| Nausea | 1 (2.8) [1] | 3 (8.1) [3] |
| Any solicited systemic AE, grade $\geq 3$ | 4 (11.1) [5] | 1 (2.7) [1] |
| **Unsolicited AEs, vaccination phase** |  |  |
| Any unsolicited AE<sup>c</sup> | 7 (19.4) [14] | 5 (13.5) [6] |
| Headache | 2 (5.6) [2] | 2 (5.4) [2] |
| Any unsolicited AE, related to study vaccination<sup>d</sup> | 1 (2.8) [2] | 1 (2.7) [1] |
| Any SAE | 0 | 0 |
| **Challenged subjects** | **N=31** | **N=32** |
| **Unsolicited AEs, post-challenge phase** |  |  |
| Any unsolicited AE<sup>c</sup> | 16 (51.6) [32] | 17 (53.1) [38] |
| Alanine aminotransferase increased | 4 (12.9) [4] | 6 (18.8) [6] |
| Aspartate aminotransferase increased | 2 (6.5) [2] | 3 (9.4) [3] |
| Post procedural contusion | 3 (9.7) [3] | 3 (9.4) [3] |
| Back pain | 2 (6.5) [3] | 0 |
| Pain in extremity | 0 | 2 (6.3) [2] |
| Headache | 0 | 2 (6.3) [2] |
| Rash maculo-papular | 0 | 2 (6.3) [2] |
| Any unsolicited AE, related to study vaccination<sup>d</sup> | 0 | 0 |
| Any unsolicited AE, related to study challenge<sup>e</sup> | 8 (22.2%) [9] | 11 (29.7%) [17] |
| Any SAE | 1 (2.8%) [1] | 1 (2.7%) [1] |
| Any SAE, related to study vaccination<sup>d</sup> | 0 | 0 |
| Any SAE, related to study challenge<sup>e</sup> | 1 (2.8%) [1] | 1 (2.7%) [1] |

AE = adverse event; N = number of participants by group that were vaccinated / challenged; n = number of participants in each group reporting the specified adverse event; RSV = respiratory syncytial virus; SAE = serious adverse event.
<sup>a</sup> Local solicited AEs included pain, erythema, swelling, induration, and pruritus on the day of vaccination and for 7 days after vaccination.
<sup>b</sup> Solicited systemic AEs included pyrexia, headache, myalgia, chills, nausea, and fatigue on the day of vaccination and for 7 days after vaccination.
<sup>c</sup> Specific unsolicited AEs are presented for events that occurred in more than one participant in either treatment group.
<sup>d</sup> Related AEs were either considered at least possibly related to study vaccine by the investigator or had missing information pertaining to relatedness.
<sup>e</sup> Related AEs were either considered at least possibly related to study challenge by the investigator or had missing information pertaining to relatedness.

## DISCUSSION

RSV remains an unconquered, ubiquitous infectious disease, and a vaccine against it has been an elusive goal through much of 60 years of research on the immunology of RSV. Human challenge trials are a unique method in clinical RSV research to study the ability of a vaccine to prevent infection under artificial, controlled exposure conditions with a viral strain that causes mild to moderate upper respiratory disease[23,24]. Human challenge trial results are not definitive, however, as research must confirm vaccine efficacy under conditions of natural infection and in the intended older adult population. Furthermore, phase 3 trials must investigate the prevention of severe disease, not merely the reduction of viral load and symptom scores, though their correlation with disease severity in natural infection has been demonstrated[25,26]. In this human challenge trial, vaccination with MVA-BN-RSV clearly resulted in lower viral load AUCs and sums of total symptom scores with similar shapes of the curves (Figure 2). The picture with infection prevention, used to measure vaccine efficacy, was somewhat more complex. The vaccine did not prevent detectable qRT-PCR confirmed infection alone, as there was little difference between the treatment groups in this regard. However, the importance of detectable RSV in the absence of symptoms or positive culture and whether it represents true RSV infection is unclear, as RNA from non-replicating virus can be detected by qRT-PCR. When infection was defined as the presence of both quantifiable viral load and clinical symptoms, vaccine efficacy was nearly 80%. When the definition of infection was further limited to the presence of live, replicating virus by culture, only 1 subject who received MVA-BN-RSV met that definition, and vaccine efficacy exceeded 85%. With this definition of infection, it appears unnecessary to use symptoms to help distinguish between infections that are clinically relevant and those that are not, and doing so did not improve point estimates of vaccine efficacy.

As useful as the human challenge model is, it is noteworthy that even in the placebo group, less than half of subjects had quantifiable virus by qRT-PCR, and less than a quarter had quantifiable culture results, despite eligibility criteria intended to select for susceptibility to RSV infection. This, along with a small sample size, made it more difficult to detect differences between the treatment groups, and vaccine efficacy confidence intervals are wide.

Scientific understanding of how immune response associates with prevention of either infection or severe disease is limited. Neutralizing antibodies to the F and G proteins are often used as a primary measure of immune response, as higher titers have been associated with reduction in disease[27,28]. Results from this study provide evidence that protective immune responses to RSV go beyond nAbs, as nAb GMTs following vaccination were lower than observed elsewhere[15,16], but high vaccine efficacy was still observed. The prognostic model for our study identified serum IgA as an important predictor of infection protection; it did not identify nAb titers, or cellular immunity against the vaccine-encoded proteins, as measured by IFN-γ ELISPOT. This was despite the fact that robust cellular responses were observed for all peptide pools, and GMSFUs remaining elevated over baseline for M2-1, N, G(A), and G(B). Prognostic models may not represent mechanistic causality, particularly in challenge trials where infection is limited to the upper respiratory tract.

The MVA-BN-RSV vaccine appears to represent a mode of action broader than other vaccine candidates focused on the production of neutralizing antibodies to the preF protein. Dependence of RSV vaccines on the activity of neutralizing antibodies against a specific epitope of one protein conformation may be risky, as such reliance may provide selective pressure for the development of mutant viruses capable of neutralizing antibody escape[29,30]. In fact, the monoclonal antibody suptavumab failed in a phase 3 clinical trial to reduce RSV hospitalizations or lower respiratory tract infections in infants, a result attributed to epitope mutations found on circulating RSV-B strains[30]. Having a vaccine that provides multiple targets for both humoral and cellular responses may protect against the consequences of such genetic pressure. The safety of MVA-BN-RSV appears to be in line with that observed with the MVA-BN vaccine, which has been administered in significant numbers both in clinical trials[31,32,33,34,35,36] and in response to the 2022 worldwide monkeypox outbreak[37]. Additionally, the MVA-BN-Filo (Mvabea) vaccine against Ebola uses the same vector[38] and has been administered to more than 200,000 people in a vaccine campaign in Rwanda[39]. No safety concerns have been identified with MVA-BN or its recombinants.

## CONCLUSIONS

MVA-BN-RSV vaccination resulted in significantly lower viral load AUC by qRT-PCR after challenge with RSV-A Memphis 37b compared to placebo. It also resulted in less infectious virus particles by culture, lower symptom scores, and estimates of vaccine efficacy in the range of 80% to 89% against infection after challenge. Humoral and cellular responses support broad immunogenicity of the vaccine. No safety issues were identified related to vaccination. Phase 3 evaluation of MVA-BN-RSV to determine clinical efficacy in an older adult population has commenced.

## Data Availability

Data produced in the present study may be available upon reasonable request to the authors

## FOOTNOTE PAGE

### Disclosures

Golam Kabir: former employee of hVIVO

Elke Jordan, Stephanie Schultz, Günter Silbernagl, Darja Schmidt, Daria Stroukova, Barbara Martin, Victoria Jenkins, Heinz Weidenthaler, Laurence De Moerlooze: current employees of Bavarian Nordic

## Acknowledgments

We want to say a special thank you to all the participants in this RSV challenge trial. Also, thanks to Bavarian Nordic colleagues and hVIVO clinical trial site investigators and personnel for their support during the conduct of the trial.

