## Supplemental methods and tables for "Decreased Viral Load, Symptom Reduction, and Prevention of Respiratory Syncytial Virus Infection with MVA-BN-RSV Vaccine"

Supplementary materials

### Inclusion Criteria

**Table of Inclusion Criteria**

| **NO** | **APPROVED INCLUSION CRITERIA** |
| --- | --- |
| To be eligible for the study, participants must meet all the following inclusion criteria: | |
| 1 | An informed consent document signed and dated by the participant and the Investigator. |
| 2 | Aged between 18 and 50 years old on the day of signing the consent form. |
| 3 | In good health with no history, or current evidence, of clinically significant medical conditions, and no clinically significant test abnormalities that will interfere with participant safety, as defined by medical history, physical examination, (including vital signs), ECG, and routine laboratory tests as determined by the Investigator. |
| 4 | A documented medical history prior to enrolment. |
| 5 | The following criteria are applicable to female participants participating in the study.   a) Females of childbearing potential must have a negative pregnancy test prior to enrolment. b) Females of non-childbearing potential: a. Post-menopausal* females; defined as having a history of amenorrhea for >12 months with no alternative medical cause, and /or by FSH level >40mLU/mL, confirmed by laboratory. b. Documented status as being surgically sterile (e.g. hysterectomy, bilateral salpingectomy and bilateral oophorectomy).   **A woman is considered of childbearing potential unless post-menopausal (defined as ≥12 months without a menstrual period) or who is permanently sterile (i.e. is at least 6*  *months post-surgical sterilization via hysterectomy or bilateral oophorectomy). Acceptable contraception methods are restricted to abstinence, intrauterine devices (IUD), intrauterine systems (IUS), licensed hormonal products or bilateral tubal ligation* |
| 6 | The following criteria apply to female and male participants:   a) Female participants of childbearing potential must use one form of highly   effective contraception. Hormonal methods must be in place from at least 2 weeks prior to the first study visit. The contraception use must continue until 28 days after the date of viral challenge. Highly effective contraception is as described below: |

| **NO** |  | **APPROVED INCLUSION CRITERIA** | |
| --- | --- | --- | --- |
| To be eligible for the study, participants must meet all the following inclusion criteria: | | | |
|  |  | a. | Established use of hormonal methods of contraception described below (for a minimum of 2 weeks prior to the first study visit). When hormonal methods of contraception are used, male are required to use a condom with a spermicide: |
|  |  |  | i. combined (oestrogen and progestogen containing) hormonal  contraception associated with inhibition of ovulation: |
|  |  |  | 1. oral |
|  |  |  | 2. intravaginal |
|  |  |  | 3. transdermal   ii. progestogen-only hormonal contraception associated with  inhibition of ovulation: |
|  |  |  | 1. oral |
|  |  |  | 2. injectable |
|  |  |  | 3. implantable |
|  |  | b. | Intrauterine device (IUD) |
|  |  | c. | Intrauterine hormone-releasing system (IUS) |
|  |  | d. | Bilateral tubal ligation |
|  |  | e. | Male sterilisation (with the appropriate post vasectomy documentation of the absence of sperm in the ejaculate) where the vasectomised male is the sole partner for that woman. |
|  |  | f. | True abstinence - sexual abstinence is considered a highly effective method only if defined as refraining from heterosexual intercourse during the entire period of risk associated with the study treatments. The reliability of sexual abstinence needs to be evaluated in relation to the duration of the clinical study and the preferred and usual lifestyle of the participant. |
|  | b) | Male participants must agree to the contraceptive requirements below from the | |
|  |  | vaccination visit and continue until 28 days after the date of Viral challenge: | |
|  |  | a. | Use a condom with a spermicide to prevent pregnancy in a female partner or to prevent exposure of any partner (male and female) to the IMP. |
|  |  | b. | Male sterilisation with the appropriate post vasectomy documentation of the absence of sperm in the ejaculate (*please note that the use of condom with spermicide will still be required to prevent partner exposure).* This applies only to males participating in the study. |
|  |  | c. | In addition, for female partners of childbearing potential, that partner must use another form of contraception such as one of the highly effective methods mentioned above for female participants. |
|  |  | d. | True abstinence - sexual abstinence is considered a highly effective method only if defined as refraining from heterosexual intercourse during the entire period of risk associated with the study treatments. The reliability of sexual abstinence needs to be evaluated in relation to the duration of the clinical study and the preferred and usual lifestyle of the participant. |

| **NO** | **APPROVED INCLUSION CRITERIA** |
| --- | --- |
| To be eligible for the study, participants must meet all the following inclusion criteria: | |
|  | c) In addition to the contraceptive requirements above, male participants must agree not to donate sperm following discharge from Quarantine until 28 days after the date of Viral Challenge/last dosing with IMP (whichever occurs last). |
| 7 | Sero-suitable to the challenge virus, as defined in the study Analytical Plan. |

### Exclusion Criteria

**Table of Exclusion Criteria**

| **NO** | **APPROVED EXCLUSION CRITERIA** |
| --- | --- |
| Participants who meet any of the following exclusion criteria will not be included in the study. | |
| **Medical History** | |
| 1 | History of, or currently active, symptoms or signs suggestive of upper or lower respiratory tract infection within 4 weeks prior to the first study visit. |
| 2 | a) Any history or evidence of any other clinically significant or currently active systemic comorbidities including psychiatric disorders (includes participants with a history of depression and/or anxiety). b) And/or other major disease that, in the opinion of the Investigator, may put the participant at undue risk, or interfere with a participant completing the study and necessary investigations (e.g., autoimmune disease or immunodeficiency).   ***Guidance***  *The following conditions apply:*   - *Participants with clinically mild atopic eczema/atopic dermatitis and clinically mild psoriasis may be included at the Investigator's discretion (e.g., if small amounts of regular topical steroids are used, no eczema in cubital fossa; moderate to large amounts of daily dermal corticosteroids is an exclusion).* - *Any rhinitis (specifically upper respiratory tract symptoms related to hay fever) which is clinically active or history of moderate to severe rhinitis, or history of seasonal allergic rhinitis likely to be active at the time of inclusion into the study and/or requiring regular nasal corticosteroids on an at least weekly basis, within 30 days of admission to Quarantine will be excluded. Participants with a history of currently inactive rhinitis (within the last 30 days) or mild rhinitis may be included at the PI’s discretion.* |

| **NO** | **hVIVO APPROVED STANDARD EXCLUSION CRITERIA** |
| --- | --- |
| Participants who meet any of the following exclusion criteria will not be included in the study. | |
|  | - *Participants with a physician diagnosed underactive thyroid who have been controlled on treatment for at least 6 months with evidence of a normal thyroid function test (TFT) can be included at the discretion of the PI.* - *Any concurrent serious illness including history of malignancy that may interfere with the aims of the study or a participant completing the study. Basal cell carcinoma within 5 years of initial diagnosis or with evidence of recurrence is also an exclusion.* - *Participants with a history of psychiatric illness including depression and/or anxiety of any severity within the last 2 years can be included if the Patient Health Questionnaire (PHQ-9) and / or the Generalised Anxiety Disorder Questionnaire (GAD-7) is less than or equal to 4. Participants with a PHQ-9 or GAD-7 score of between 5 and 9 may be included following consultation with a Senior Physician (Clinical Lead for Screening) who may advise further consultation with the PI.* - *Participants reporting physician diagnosed migraine can be included provided there are no associated neurological symptoms such as hemiplegia or visual loss. Cluster headache/migraine or prophylactic treatment for migraine is an exclusion.* - *Participants with physician diagnosed mild Irritable Bowel Syndrome (IBS) not requiring regular treatment can be included at the discretion of the PI.* |
| 3 | Participants who have smoked ≥10 pack years at any time **[10 pack years is equivalent to one pack of 20 cigarettes a day for 10 years].** |
| 4 | A total body weight ≤50 kg or Body Mass Index (BMI) ≤18 kg/m2 or ≥35kg/m^2^. |
| 5 | Females who:   a) Are breastfeeding, or b) Have been pregnant within 6 months prior to the study. |
| 6 | History of anaphylaxis-and/or a history of severe allergic reaction or significant intolerance to any food or drug or vaccine, as assessed by the PI. |
| 7 | Venous access deemed inadequate for the phlebotomy and cannulation demands of the study. |
| 8 | a) Any significant abnormality altering the anatomy of the nose in a substantial way or nasopharynx that may interfere with the aims of the study and in particular any of the nasal assessments or viral challenge (historical nasal polyps can be included, but large nasal polyps causing current and significant symptoms and/or requiring regular treatments in the last month will be excluded). b) Any clinically significant history of epistaxis (large nosebleeds) within the last 3 months of the first study visit and/or history of being hospitalized due to epistaxis on any previous occasion. c) Any nasal or sinus surgery within 3 months of the first study visit. |
| **Prior or Concomitant Medications and Assessments** | |

| **NO** | **hVIVO APPROVED STANDARD EXCLUSION CRITERIA** |
| --- | --- |
| Participants who meet any of the following exclusion criteria will not be included in the study. | |
| 9 | Unless medically necessary (e.g. during an outbreak or pandemic situation) and at the PI’s discretion   a) Evidence of vaccinations within the 4 weeks prior to the planned date of viral challenge/first dosing with IMP (whichever occurs first). b) Intention to receive any vaccination(s) before the last day of Follow-up. (NB. No travel restrictions will apply after the Day 28 Follow-up visit). |
| 10 | Receipt of blood or blood products, or loss (including blood donations) of 550 mL or more of blood during the 3 months prior to the planned date of viral challenge/first dosing with IMP (whichever occurs first) or planned during the 2 months after the viral challenge. |
| 11 | a) Receipt of any investigational drug within 3 months prior to the planned date of viral challenge/first dosing with IMP (whichever occurs first). b) Previous vaccination with any licensed or investigational RSV vaccine before enrolment into the study. c) Receipt of three or more investigational drugs within the previous 12 months prior to the planned date of viral challenge/first dosing with IMP (whichever occurs first). d) Prior inoculation with a virus from the same virus-family as the challenge virus. e) Prior participation in another human viral challenge study with a respiratory virus in the preceding 3 months, taken from the date of viral challenge in the previous study to the date of expected viral challenge in this study. f) Receipt of treatment with immunosuppressive therapy. |
| 12 | a) Confirmed positive test for drugs of abuse and cotinine on first study visit. One repeat test allowed at PI discretion. b) History or presence of alcohol addiction, or excessive use of alcohol (weekly intake in excess of 28 units alcohol; 1 unit being a half glass of beer, a small glass of wine or a measure of spirits), or excessive consumption of xanthine containing substances (e.g. daily intake in excess of 5 cups of caffeinated drinks e.g. coffee, tea, cola). |
| 13 | A forced expiratory volume in 1 second (FEV1) <80%. |
| 14 | Positive human immunodeficiency virus (HIV), active hepatitis A (HAV), B (HBV), or C (HCV) test. |
| **Other** | |
| 15 | Those employed or immediate relatives of those employed at hVIVO, Bavarian Nordic or any vendor. |
| 16 | Any other finding that, in the opinion of the Investigator, deems the Participant unsuitable for the study. |

### Symptom Diary Cards used During Quarantine Stay

Participants reported and assessed the severity of any challenge virus-related signs and symptoms three times/day until to planned discharge from Quarantine, at the same time each day (±1 hour), using the hVIVO Symptom Diary Card. This information was collected using a paper form.

The following symptoms in the 13-item symptom questionnaire were graded on a scale of 0-3 (Grade 0: No symptoms; Grade 1: just noticeable; Grade 2: clearly bothersome from time to time but does not interfere with me doing my normal daily activities; Grade 3: Quite bothersome most or all of the time, and it stops me participating in activities). Shortness of breath and Wheeze had an additional Grade 4: Symptoms at rest.

- Runny nose
- Stuffy nose
- Sneezing
- Sore throat
- Earache
- Malaise/tiredness
- Headache
- Muscle and/or joint ache
- Chilliness/Feverishness
- Cough
- Chest tightness
- Shortness of breath
- Wheeze

### Reverse Transcription Polymerase Chain Reaction Assay

**Pan Real-Time Quantitative Reverse Transcription Polymerase Chain Reaction (RT-qPCR) Assay for the Detection of Respiratory Syncytial Virus (RSV) RNA**

A RT-qPCR using a primer and probe set designed on a universally conserved RSV sequence was validated and used for the detection and quantification of RSV‑A Memphis 37b.

RSV RNA was extracted from samples using a Qiagen DSP Virus/Pathogen Midi kit on the QIASymphony SP module (Qiagen). The PCR reactions were loaded in triplicate by the QIASymphony AS module into Qiagen OneStep QuantiFAST mastermix containing a RSV Taqman hydrolysis probe and PCR primers targeting the RSV genome. The one-step RT-qPCR reaction was performed using a ViiA7 thermocycler (Applied Biosystems). The quantity of viral RNA was determined by the comparison of cycle threshold (Ct) values to a calibration standard curve prepared from in vitro generated nucleic acid copies of the target RSV gene sequence.

The LLOQ was defined as a Ct value of 33.9. The ULOQ was defined as a Ct value of 16.1.

### Viral Culture Methods

Infectious virus in nasal washes was quantified using a validated virus plaque assay. Human epithelial type 2 (HEp-2) cells were seeded into 24- well plates and incubated at 37˚C (±2˚C), 5% CO2, until 80 – 90% confluent on the day of infection. Test samples were prepared at 4 dilutions. The cell culture medium was removed from the HEp-2 cells and the monolayer inoculated by adding each dilution of test sample in triplicate. The assay plates were incubated for 1 hour at 37˚C (±2˚C), 5% CO2. After incubation, pre-warmed methyl cellulose media was added and the plates incubated for 6 days at 37˚C (±2˚C), 5% CO2. The methylcellulose media was removed and the cells fixed with buffered formaldehyde. The cell monolayer was first stained with haematoxylin solution followed by eosin Y solution; the cell monolayer was then washed with water and the plate dried before the presence of plaques was determined by light microscopy. Only wells containing ≤50 plaques were counted and used in the quantification of the titer. The virus titer, in log10 Plaque Forming Units (PFU)/mL, was calculated for each dilution from the average number of plaques obtained for the three replicate wells. The test sample final titer was defined as the mean titer of all valid dilutions.

Table S1:	Demographics and Baseline Characteristics

| Description | Statistics | MVA-BN-RSV (N=36) | Placebo (N=37) | Total (N=73) |
| --- | --- | --- | --- | --- |
| Age (years) | Mean (SD) | 26.1 (5.2) | 25.7 (6.5) | 25.9 (5.9) |
|  | Median | 26.5 | 25.0 | 25.0 |
|  | Min, Max | 18, 42 | 18, 50 | 18, 50 |
| Sex: n (%) | Female | 13 (36.1) | 17 (45.9) | 30 (41.1) |
|  | Male | 23 (63.9) | 20 (54.1) | 43 (58.9) |
| Race: n (%) | White | 33 (91.7) | 33 (89.2) | 66 (90.4) |
|  | Asian | 1 (2.8) | 2 (5.4) | 3 (4.1) |
|  | Multiple | 2 (5.6) | 2 (5.4) | 4 (5.5) |
| BMI (kg/m^2^) at screening | Mean (SD) | 24.48 (3.09) | 24.94 (3.65) | 24.71 (3.37) |
|  | Median | 25.05 | 24.80 | 24.90 |
|  | Min, Max | 18.3, 30.1 | 18.9, 34.1 | 18.3, 34.1 |

BMI = body mass index; kg = kilograms; m = meters; N = number of participants in the specified group; n = number of participants within the specified group contributing to statistic; SD = standard deviation; % = percentage based on N.

Descriptive analyses performed on the safety population analysis set.

Table S2:	RSV-specific Humoral Responses

|  | IgA^a^ | | IgG^a^ | | nAb^b^, RSV-A | | nAb^b^, RSV-B | |
| --- | --- | --- | --- | --- | --- | --- | --- | --- |
| Time Point  Statistic | MVA‑BN‑RSV  **(N=30)** | Placebo  **(N=31)** | MVA‑BN‑RSV  (N=30) | Placebo  (N=31) | MVA‑BN‑RSV  (N=30) | Placebo  (N=31) | MVA‑BN‑RSV  (N=30) | Placebo  (N=31) |
| Day -28 | | | | | | | | |
| n | 30 | 31 | 30 | 31 | 30 | 31 | 30 | 31 |
| GMT | 264.3 | 214.9 | 815.4 | 711.6 | 287.7 | 263.8 | 174.8 | 204.9 |
| 95% CI | (206.1, 338.9) | (139.4, 331.2) | (673.2, 987.6) | (585.5, 864.8) | (218.9, 378.0) | (192.9, 360.9) | (117.5, 260.2) | (141.4, 296.9) |
| **Day -14 (14 Days After Vaccination)** | | | | | | | | |
| n | 30 | 31 | 30 | 31 | 30 | 31 | 30 | 31 |
| GMT | 994.0 | 197.4 | 3268.2 | 707.3 | 600.2 | 335.3 | 277.8 | 177.2 |
| 95% CI | (728.8, 1355.8) | (129.3; 301.3) | (2651.2, 4028.9) | (580.6, 861.8) | (473.0, 761.5) | (288.2, 390.0) | (198.6, 388.5) | (122.7, 255.9) |
| **Day -2 (Quarantine Admission)** | | | | | | | | |
| n | 30 | 31 | 30 | 31 | 30 | 31 | 30 | 31 |
| GMT | 540.5 | 202.1 | 3100.6 | 737.9 | 494.8 | 276.6 | 242.0 | 181.0 |
| 95% CI | (396.5, 736.9) | (136.7, 298.6) | (2499.3, 3846.6) | (596.9, 912.1) | (394.7, 620.3) | (201.0, 380.5) | (163.2, 358.7) | (124.4, 263.3) |
| **Day 5** | | | | | | | | |
| n | 29 | 31 | 29 | 31 | 29 | 31 | 29 | 31 |
| GMT | 544.0 | 220.3 | 2726.4 | 713.2 | 427.5 | 327.4 | 233.8 | 188.2 |
| 95% CI | (396.3; 746.9) | (149.2; 325.4) | (2240.4, 3317.8) | (586.8, 867.0) | (344.9, 529.9) | (280.3, 382.3) | (158.8, 344.1) | (134.3, 263.7) |
| **Day 10** | | | | | | | | |
| n | 30 | 31 | 30 | 31 | 30 | 31 | 30 | 31 |
| GMT | 504.9 | 398.5 | 2888.2 | 1139.4 | 443.5 | 433.5 | 247.6 | 291.0 |
| 95% CI | (370.7; 687.5) | (274.9; 577.7) | (2437.8, 3421.7) | (894.5, 1451.5) | (341.1, 576.5) | (313.1, 600.2) | (166.7, 367.8) | (200.5, 422.4) |
| **Day 28** | | | | | | | | |
| n | 29 | 31 | 29 | 31 | 29 | 31 | 29 | 31 |
| GMT | 552.5 | 561.2 | 2871.6 | 1762.7 | 519.4 | 751.2 | 246.5 | 393.9 |
| 95% CI | (400.4; 762.5) | (407.9; 772.1) | (2432.8, 3389.6) | (1314.7, 2363.4) | (410.3, 657.5) | (523.6, 1077.8) | (168.6, 360.4) | (265.4, 584.6) |
| **Day 155** | | | | | | | | |
| n | 29 | 28 | 29 | 28 | 29 | 28 | 29 | 28 |
| GMT | 549.1 | 414.2 | 1961.0 | 1203.7 | 441.6 | 475.2 | 235.9 | 253.0 |
| 95% CI | (409.3; 736.6) | (288.0; 595.6) | (1608.6, 2390.6) | (958.0, 1512.4) | (338.3, 576.3) | (363.1, 621.8) | (158.0, 352.4) | (171.3, 373.7) |

CI = confidence interval; ELISA = enzyme-linked immunosorbent assay; GMT = geometric mean titer; IgA = immunoglobulin A; IgG = immunoglobulin G; IU = infectious units; N = number of subjects in specified group; n = number of subjects within specified group contributing to statistic; nAb = neutralizing antibodies; PRNT = plaque reduction neutralization test; RSV = respiratory syncytial virus.

^a^	Titer determined by ELISA.

^b^	Titer determined by PRNT, standardized to World Health Organization standard as IU/mL.

The results <LLOQ are considered by using ½ LLOQ for the calculation of statistics of this parameter.

Analyses were performed on the per protocol analysis set.

Table S3:	Cellular Immune Response: Enumeration of Interferon-γ-producing Peripheral Blood Mononuclear Cells

|  | IFN-γ ELISPOT Pool F | | IFN-γ ELISPOT Pool GA | | IFN-γ ELISPOT Pool GB | | IFN-γ ELISPOT Pool M2 | | IFN-γ ELISPOT Pool N | | IFN-γ ELISPOT Pool RSV | |
| --- | --- | --- | --- | --- | --- | --- | --- | --- | --- | --- | --- | --- |
| Time  Statistic | MVA-BN-RSV (N=30) | Placebo (N=31) | MVA-BN-RSV (N=30) | Placebo (N=31) | MVA-BN-RSV (N=30) | Placebo (N=31) | MVA-BN-RSV (N=30) | Placebo (N=31) | MVA-BN-RSV (N=30) | Placebo (N=31) | MVA-BN-RSV (N=30) | Placebo (N=31) |
| **Day -28** | | | | | | | | | | | | |
| n | 19 | 24 | 19 | 24 | 19 | 24 | 19 | 24 | 19 | 24 | 19 | 24 |
| GMSFU | 365.2 | 298.6 | 160.7 | 142.3 | 151.6 | 133.2 | 49.0 | 50.3 | 204.1 | 195.8 | 413.7 | 524.8 |
| 95% CI | (241.5, 552.3) | (189.9, 469.5) | (92.8, 278.4) | (81.1, 249.7) | (93.0, 247.2) | (73.2, 242.5) | (32.78, 73.2) | (33.6, 75.5) | (128.3, 324.7) | (111.8, 343.0) | (246.6, 694.1) | (397.5, 692.9) |
| **Day -21 (7 Days After Vaccination)** | | | | | | | | | | | | |
| n | 28 | 27 | 28 | 27 | 28 | 27 | 28 | 27 | 28 | 27 | 28 | 27 |
| GMSFU | 783.1 | 260.3 | 483.1 | 113.6 | 435.0 | 102.8 | 320.5 | 48.1 | 731.0 | 222.4 | 854.5 | 399.6 |
| 95% CI | (576.1, 1064.5) | (176.0, 384.9) | (309.6, 753.8) | (67.3, 191.7) | (254.3, 744.0) | (61.1, 172.9) | (209.9, 489.4) | (33.7, 68.6) | (510.7, 1046.3) | (145.1, 341.0) | (588.2, 1241.4) | (273.8, 583.2) |
| **Day -2 to 0 (Quarantine Admission, Prior to Challenge)** | | | | | | | | | | | | |
| n | 23 | 24 | 23 | 24 | 23 | 24 | 23 | 24 | 23 | 24 | 23 | 24 |
| GMSFU | 637.3 | 271.7 | 408.5 | 152.0 | 336.7 | 118.0 | 220.6 | 44.5 | 565.2 | 258.7 | 1014.5 | 455.1 |
| 95% CI | (498.0, 815.6) | (175.6, 420.3) | (284.90, 585.63) | (93.2, 247.9) | (204.2, 555.3) | (69.5, 200.4) | (130.3, 373.7) | (30.1, 65.7) | (446.9, 714.8) | (170.5, 392.5) | (813.1, 1265.8) | (330.9, 626.0) |
| **Day 5** | | | | | | | | | | | | |
| n | 25 | 25 | 25 | 25 | 25 | 25 | 25 | 25 | 25 | 25 | 25 | 25 |
| GMSFU | 464.2 | 187.6 | 334.2 | 83.5 | 299.8 | 79.4 | 175.8 | 42.4 | 405.5 | 138.8 | 689.4 | 302.3 |
| 95% CI | (300.2, 717.8) | (116.5, 302.2) | (227.3, 491.33) | (51.3, 135.8) | (203.0, 442.7) | (47.9, 131.6) | (112.0, 276.1) | (28.9, 62.3) | (254.8, 645.3) | (83.5, 230.7) | (436.7, 1088.1) | (182.4, 501.1) |
| **Day 10** | | | | | | | | | | | | |
| n | 24 | 26 | 24 | 26 | 24 | 26 | 24 | 26 | 24 | 26 | 24 | 26 |
| GMSFU | 545.2 | 622.4 | 481.9 | 392.2 | 384.1 | 300.1 | 232.8 | 202.6 | 450.5 | 491.3 | 757.4 | 878.2 |
| 95% CI | (355.3, 836.5) | (440.2, 880.0) | (341.5, 679.9) | (230.1, 668.4) | (255.3, 577.8) | (172.9, 520.9) | (150.4, 360.3) | (113.3, 362.3) | (291.2, 697.1) | (353.0, 683.7) | (475.6, 1206.3) | (601.9, 1281.5) |
| **Day 28** | | | | | | | | | | | | |
| n | 24 | 23 | 24 | 23 | 24 | 23 | 24 | 23 | 24 | 23 | 24 | 23 |
| GMSFU | 451.5 | 394.3 | 298.2 | 195.4 | 263.6 | 150.2 | 180.6 | 111.9 | 455.7 | 311.1 | 691.6 | 507.4 |
| 95% CI | (293.8, 693.9) | (267.9, 580.2) | (194.9, 456.3) | (109.3, 349.2) | (164.2, 423.2) | (79.5, 283.6) | (120.0, 271.8) | (68.1, 183.9) | (325.1, 638.8) | (186.9, 517.8) | (488.3, 979.7) | (300.1, 858.0) |
| **Day 155** | | | | | | | | | | | | |
| n | 22 | 27 | 22 | 27 | 22 | 27 | 22 | 27 | 22 | 27 | 22 | 27 |
| GMSFU | 347.7 | 369.5 | 263.7 | 175.4 | 226.6 | 142.3 | 172.9 | 61.4 | 413.2 | 241.6 | 553.2 | 514.3 |
| 95% CI | (219.2, 551.5) | (265.4, 514.6) | (171.7, 405.0) | (105.7, 291.0) | (141.0, 364.3) | (84.4, 239.8) | (112.1, 266.6) | (41.2, 91.6) | (301.3, 566.8) | (156.3, 373.5) | (338.7, 903.5) | (343.7, 769.7) |

CI = confidence interval; ELISPOT = enzyme-linked immune absorbent spot; GMSFU = geometric mean spot forming unit; IFN-γ = interferon-gamma; LLOQ = lower limit of quantification; N = number of participants in a specified group; n = number of participants within a specified group contributing to the statistic.

The results <LLOQ are considered by using ½LLOQ for the calculation of statistics for the various pool.
